# Reducing Stigma and Bias in Perinatal Substance Use Care: A Training for Obstetric and Neonatal Providers

**DOI:** 10.1101/2025.06.17.25329801

**Authors:** Karli S Swenson, Katie Breen

**Affiliations:** Colorado Perinatal Care Quality Collaborative

## Abstract

**Background:** Accidental overdose is the second leading cause of death among pregnant and postpartum individuals in Colorado, with substance use disorders (SUD) contributing significantly to maternal morbidity. Stigma and bias from healthcare providers exacerbate these challenges, leading to suboptimal care and reduced access to essential resources. Despite extensive documentation of negative attitudes toward pregnant individuals with SUD, few interventions aim to reduce such biases in healthcare settings.

**Objective:** To evaluate the impact of a 90-minute live, virtual or in person training on reducing stigma and bias among healthcare providers caring for patients with perinatal SUD.

**Methods:** This pre-post intervention study enrolled healthcare professionals, including obstetric and neonatal nurses, obstetricians, midwives, and nurse educators, who provide care to perinatal patients. Participants completed a training developed by the Colorado Perinatal Care Quality Collaborative (CPCQC) in partnership with experts with lived experience of perinatal SUD (“lived-experience experts”) from HardBeauty. The training focused on evidence-based education, stigma and bias reduction, and patient-centered strategies informed by lived experiences. Outcomes were assessed, a modified pre-post-training knowledge and comfortability survey using Likert scales and open response questions, and post-training qualitative feedback.

**Results:** Participants (n=549) demonstrated statistically significant improvements in response scores (p < 0.05), indicating reduced stigma and bias toward perinatal patients with SUD. Qualitative feedback highlighted the training’s relevance and impact, with participants emphasizing the value of integrating lived experiences into educational initiatives.

**Conclusions:** This study demonstrates the effectiveness of a brief, virtual training in reducing stigma and bias among healthcare providers caring for patients with perinatal SUD. The findings underscore the importance of incorporating lived-experience expertise into training programs to promote equitable and compassionate care. Future research should explore long-term impacts on clinical practice and patient outcomes.

## Introduction

Accidental overdose is the second leading cause of death among pregnant and postpartum individuals in Colorado, underscoring an urgent and under-addressed public health crisis.^1^ The perinatal period represents a time of both heightened vulnerability and opportunity, yet substance use remains a significant contributor to maternal morbidity, with profound implications for maternal and neonatal outcomes.^2^ Despite the availability of pharmacologic, social, and behavioral health interventions, systemic barriers, including stigma and bias, impede effective care delivery.^3^

Substantial evidence demonstrates that pregnant and postpartum individuals with substance use disorders (SUD) encounter stigma and bias from healthcare providers, particularly within obstetrics and labor and delivery units.^4^ These attitudes result in suboptimal clinical care, eroding trust and preventing patients from accessing evidence-based resources critical to their health and recovery.^3-5^ The ripple effects of stigma extend beyond the individual, exacerbating disparities in maternal and child health outcomes and increasing the burden on healthcare systems.^6^

While the literature has extensively documented negative attitudes and behaviors among healthcare professionals toward perinatal patients with SUD,^3-5^ interventions designed to reduce these biases remain sparse. Existing training programs often lack a focus on the lived experiences of individuals with SUD or fail to integrate actionable strategies for improving clinical care and resource access.^7-14^ This gap underscores an unmet need for scalable, evidence-based interventions tailored to the unique challenges of perinatal substance use.

This study introduces a novel approach: a 90-minute, live, virtual or in person training developed by the Colorado Perinatal Care Quality Collaborative (CPCQC) in partnership with HardBeauty, an organization led by experts with lived experience of perinatal SUD. By integrating real-world insights and leveraging a multidisciplinary framework, this training aims to reduce stigma and bias among healthcare providers, equipping them with the tools to deliver equitable and compassionate care. To our knowledge, this is one of the first studies to rigorously evaluate the impact of such training in a perinatal care setting, providing a model for replication and scale across diverse healthcare systems.

## Methods

### Study Design

This study utilized a modified pre-post intervention design to evaluate the impact of a 90-minute live, virtual training on reducing stigma and bias among healthcare providers caring for patients with perinatal substance use disorders (SUD). The training was developed by the Colorado Perinatal Care Quality Collaborative (CPCQC) in collaboration with HardBeauty, an organization specializing in lived-experience expertise of SUD. After initial development of the training, peer support experience expanded to include the University of Colorado College of Nursing (CU CON) Peer Recovery Doula program.

### Participants

Participants included healthcare professionals providing care to perinatal and neonatal patients, including nurses, obstetricians, pediatricians, midwives, social workers, lactation consultants, and nurse educators. Recruitment was conducted through CPCQC member organizations participating in the Turning the Tide perinatal substance use quality improvement initiative. Hospital teams must have either labor and delivery (L&D) or neonatal intensive care units (NICUs) on site to be eligible. Participation was encouraged but voluntary, any questions could be skipped, and all participants provided informed consent before completing the survey. Of the 1,454 healthcare providers who completed the training between 2023 and 2025, 598 opened the post-training survey and 549 completed at least 80% of the survey items. Only these 549 responses were included in the final analysis to ensure data completeness. Because demographic information was only collected from participants who completed the survey, we were unable to assess differences between responders and non-responders. As such, we could not formally evaluate the presence or impact of nonresponse bias. All quantitative analyses used a complete case approach, excluding individuals with substantial missing data (<80% survey completion). While this strategy preserved data integrity, it may limit the generalizability of findings to the broader population of training participants.

### Training Intervention

The 90-minute training was delivered live and virtually via a secure video conferencing platform (Zoom or Microsoft Teams). The curriculum included three core components:

1. Education on perinatal SUD: Evidence-based content on social determinants of health and trauma-informed care in perinatal substance use, highlighting the trajectories of individuals who develop substance use disorders.
2. Toolkit on caring for patients with perinatal SUD: 10-step toolkit on how appropriate care provided by units and individual staff can directly benefit patients with perinatal SUD.
3. Integration of Lived Experience: Humanizing narratives and actionable clinical strategies provided by HardBeauty or Peer Recovery Doula experts, emphasizing patient-centered approaches to care.

Interactive components, such as polls, case studies, and breakout room discussions, were incorporated to promote engagement and application of concepts.

### Evaluation

Evaluation followed best practices for training assessment, aligning with Level 1 (reaction),s Level 2 (learning), and elements of Level 3 (behavioral intention) of the Kirkpatrick Model for evaluating training effectiveness. Participants completed an anonymous survey immediately before and after the training. The survey included 4-point Likert-scale questions assessing changes in attitudes, knowledge, and comfort across multiple domains. Pre-training items assessed stigma-related beliefs, empathy, and perceived knowledge about perinatal SUD and maternal health disparities. Both pre- and post-training surveys evaluated comfort with clinical and interpersonal scenarios, including screening for substance use, engaging in nonjudgmental counseling, initiating harm reduction conversations, and intervening in stigmatizing peer behaviors. Post-training items captured changes in self-reported knowledge, confidence in applying learned content, and intention to share information with colleagues. Open-ended questions solicited qualitative feedback on perceived barriers to care, reflections on the impact of lived-experience experts, and interest in future engagement with peer recovery coaches. Demographic questions captured participants’ hospital affiliation, education level, race/ethnicity, unit type, duration of service on the unit, and previous training or exposure to perinatal SUD care. Data were analyzed descriptively and thematically, and paired pre- and post-training responses were compared to evaluate changes in attitudes and comfort levels. A follow-up survey was conducted six months post-training to assess retention of knowledge and sustained changes in attitudes. All surveys were distributed electronically via SurveyMonkey and consent to participate in research was collected before survey initiation. Likert scales were used on a 1-4 scale with 1 being strongly disagree/very uncomfortable and 4 being strongly agree/very comfortable..

### Analysis

Pre- and post-training survey responses were compared using Wilcoxon signed-rank tests, as data were non-normally distributed and paired by respondent. To evaluate differences between virtual and in-person training modalities, Wilcoxon rank-sum (Mann-Whitney U) tests were used to compare independent groups. Changes in median scores were reported alongside interquartile ranges (IQR) to reflect the magnitude and direction of observed effects. Open-ended survey responses were analyzed using inductive thematic analysis to identify recurring patterns and key concepts. One primary coder reviewed all responses and applied initial thematic labels. Themes were iteratively refined through consensus discussion, and illustrative quotes were selected to highlight the depth and variability of participant reflections. Thematic domains were derived directly from the data rather than imposed a priori, allowing for authentic insights to emerge across participant roles and settings.^15^

## Ethical Considerations

The study protocol was deemed quality improvement by the University of Colorado Anschutz Medical Campus (IRB approval #25-0659), though consent to participate in research was collected, nonetheless.

Participants’ data were anonymized, and confidentiality was maintained throughout the study.

## Results

In total, 1,454 staff completed the training, with 111 in late 2023, 685 in 2024, and 658 in 2025. 29 hospitals, one birth center, and 3 nonprofit nurse-family partnership organizations hosted the training. The combined 1,343 participants from 2024 and 2025 were eligible to participate in the finalized program evaluation. Of those, 598 opened the survey and 549 completed >80% of the questions. 443 completed the final section of demographics. The survey took 14 minutes on average to complete. The majority of attendees (78.8%) were nurses with a Bachelor of Science in Nursing. Most identified as white (82.4%) and worked in labor and delivery (36.4%), postpartum/mom-baby units (39.8%), or the NICU (37.6%) (Table 1). Attendees varied in experience, ranging from early-career professionals with 0-5 years of experience (34.7%) to those with over 20 years in practice (18.5%). Regarding background knowledge on perinatal substance use disorders (SUD), 269 (60.7%) attendees reported on-the-job training, and 197 (44.5%) had completed online learning modules. Other common responses included that SUD was mentioned in their education but not in sufficient depth to prepare them for patient care (169, 38.1%), and having personal experience with friends or family members who have substance use

**Table 1.**
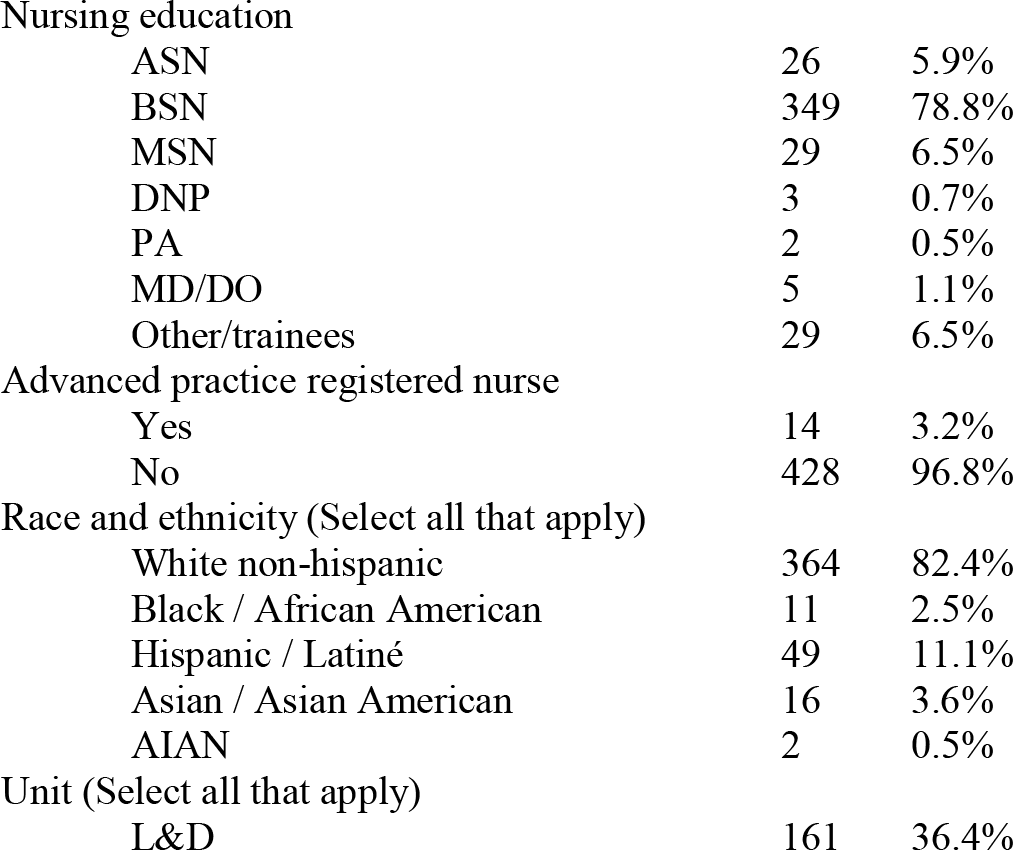
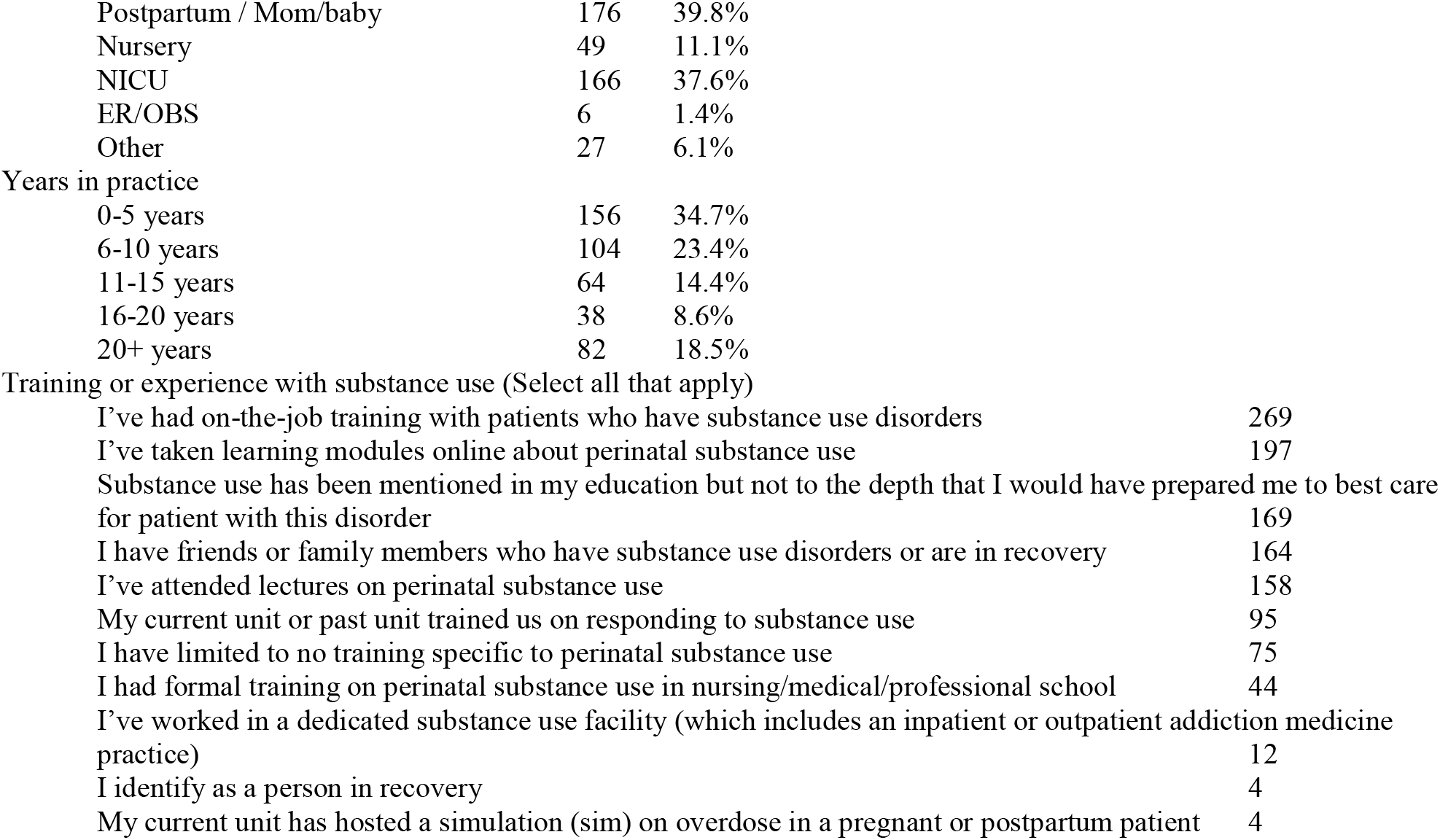
Demographics of training attendees.

|  |  |  |
| --- | --- | --- |
| Nursing education |  |  |
| ASN | 26 | 5.9% |
| BSN | 349 | 78.8% |
| MSN | 29 | 6.5% |
| DNP | 3 | 0.7% |
| PA | 2 | 0.5% |
| MD/DO | 5 | 1.1% |
| Other/trainees | 29 | 6.5% |
| Advanced practice registered nurse |  |  |
| Yes | 14 | 3.2% |
| No | 428 | 96.8% |
| Race and ethnicity (Select all that apply) |  |  |
| White non-hispanic | 364 | 82.4% |
| Black / African American | 11 | 2.5% |
| Hispanic / Latiné | 49 | 11.1% |
| Asian / Asian American | 16 | 3.6% |
| AIAN | 2 | 0.5% |
| Unit (Select all that apply) |  |  |
| L&D | 161 | 36.4% |
| Postpartum / Mom/baby | 176 | 39.8% |
| Nursery | 49 | 11.1% |
| NICU | 166 | 37.6% |
| ER/OBS | 6 | 1.4% |
| Other | 27 | 6.1% |
| Years in practice |  |  |
| 0-5 years | 156 | 34.7% |
| 6-10 years | 104 | 23.4% |
| 11-15 years | 64 | 14.4% |
| 16-20 years | 38 | 8.6% |
| 20+ years | 82 | 18.5% |
| Training or experience with substance use (Select all that apply) |  |  |
| I've had on-the-job training with patients who have substance use disorders |  | 269 |
| I've taken learning modules online about perinatal substance use |  | 197 |
| Substance use has been mentioned in my education but not to the depth that I would have prepared me to best care for patient with this disorder |  | 169 |
| I have friends or family members who have substance use disorders or are in recovery |  | 164 |
| I've attended lectures on perinatal substance use |  | 158 |
| My current unit or past unit trained us on responding to substance use |  | 95 |
| I have limited to no training specific to perinatal substance use |  | 75 |
| I had formal training on perinatal substance use in nursing/medical/professional school |  | 44 |
| I've worked in a dedicated substance use facility (which includes an inpatient or outpatient addiction medicine practice) |  | 12 |
| I identify as a person in recovery |  | 4 |
| My current unit has hosted a simulation (sim) on overdose in a pregnant or postpartum patient |  | 4 |

To gauge baseline attitudes and biases towards pregnant patients with SUDs, we asked a variety of questions related to the competency and experience with patients with SUDs. While staff strongly disagreed that patients with SUD “cannot be helped,” and strongly agreed patients with SUD often have experiences of physical, sexual, or emotional trauma, some questions had a greater degree of variability in responses. Staff generally agreed (median, at “somewhat agree”) that persons with SUD could quit using substances “if they really wanted to,” “if they’re provided the needed supports and resources,” and that SUDs often develop following prescription of pain medication. Staff generally disagreed (median, at “somewhat disagree”) that pregnant patients with with SUDs are “irresponsible and negligent,” that they as professionals generally “feel burnt out” or “frustrated” working with patients with SUD, and that “criminal prosecution for fetal abuse following perinatal SUD protects the child.” In general, these responses are sympathetic towards this patient population.

Participants demonstrated statistically significant increases in knowledge, empathy, and trauma-informed perspectives following the training (Table 3). Before the session, self-reported knowledge of key topics, such as the prevalence of substance use in pregnancy (median = 3.0 [2.0, 3.0]) and the contribution of overdose and suicide to maternal mortality in Colorado (2.0 [1.0, 3.0]), was moderate to low. Post-training responses reflected substantial gains, with median scores rising to 4.0 [4.0, 4.0] for both items (p = 1.15e-73 and 2.04e-76, respectively). Similarly, participants reported increased understanding of racial disparities in maternal outcomes (pre: 3.0 [2.0, 4.0]; post: 4.0 [4.0, 4.0]; p = 1.28e-59) and greater awareness that patients with substance use disorders often fear child welfare involvement (pre: 4.0 [3.0, 4.0]; post: 4.0 [4.0, 4.0]; p = 4.76e-34).

**Table 2.**
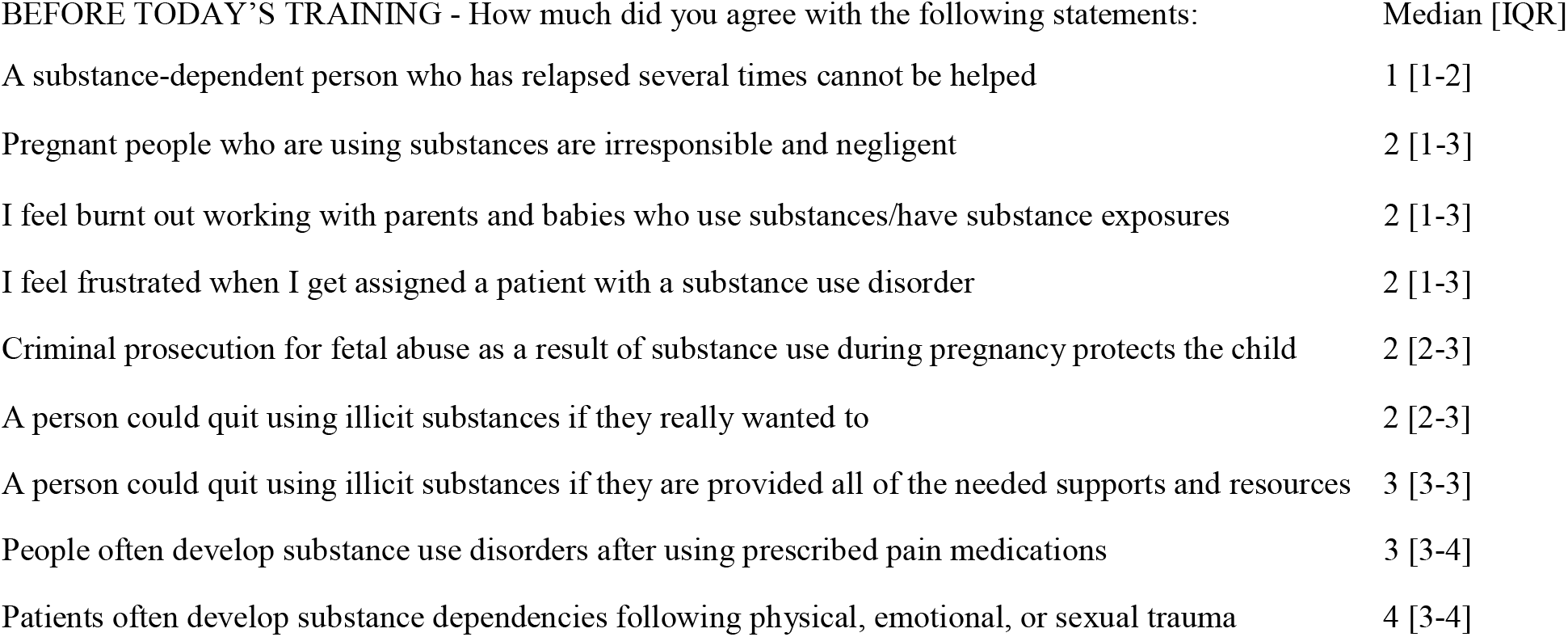
Baseline measures.

| BEFORE TODAY’S TRAINING - How much did you agree with the following statements: | Median [IQR] |
| --- | --- |
| A substance-dependent person who has relapsed several times cannot be helped | 1 [1-2] |
| Pregnant people who are using substances are irresponsible and negligent | 2 [1-3] |
| I feel burnt out working with parents and babies who use substances/have substance exposures | 2 [1-3] |
| I feel frustrated when I get assigned a patient with a substance use disorder | 2 [1-3] |
| Criminal prosecution for fetal abuse as a result of substance use during pregnancy protects the child | 2 [2-3] |
| A person could quit using illicit substances if they really wanted to | 2 [2-3] |
| A person could quit using illicit substances if they are provided all of the needed supports and resources | 3 [3-3] |
| People often develop substance use disorders after using prescribed pain medications | 3 [3-4] |
| Patients often develop substance dependencies following physical, emotional, or sexual trauma | 4 [3-4] |

**Table 3.** Knowledge of perinatal SUD.

| How much did you agree with the following statements: | Before training: | After training: | p value |
| --- | --- | --- | --- |
| I knew how prevalent substance use was in pregnancy | 3.0 [2.0, 3.0] | 4.0 [4.0, 4.0] | 1.15E-73 |
| I knew how much overdose and suicide contribute to maternal mortality in Colorado | 2.0 [1.0, 3.0] | 4.0 [4.0, 4.0] | 2.04E-76 |
| I knew the disparities in maternal morbidity and mortality for Native and Black populations in Colorado | 3.0 [2.0, 4.0] | 4.0 [4.0, 4.0] | 1.28E-59 |
| I knew that parents with substance use disorders may fear CPS due to the threat of taking their baby away | 4.0 [3.0, 4.0] | 4.0 [4.0, 4.0] | 4.76E-34 |
| I have thought about the circumstances of a patient's life story when told they have a substance use disorder while pregnant | 3.0 [3.0, 4.0] | 4.0 [4.0, 4.0] | 2.22E-57 |
| I felt empathy for pregnant/postpartum people with substance use disorders | 3.0 [3.0, 4.0] | 4.0 [4.0, 4.0] | 1.37E-50 |
| I viewed addiction as a preventable and treatable disease that a patient is facing | 3.0 [3.0, 3.0] | 4.0 [3.0, 4.0] | 6.19E-49 |

Shifts in empathy and mindset were also observed. Agreement with the statements “I have thought about the circumstances of a patient’s life story…” and “I view addiction as a preventable and treatable disease…” significantly increased following the training (pre: 3.0 [3.0, 4.0] and 3.0 [2.0, 3.0]; post: 4.0 [4.0, 4.0] for both; p = 2.22e-57 and 2.37e-52, respectively). Notably, even items rated relatively highly at baseline, such as empathy for perinatal patients with substance use disorders (3.0 [3.0, 3.0]), demonstrated meaningful increases post-training (4.0 [4.0, 4.0]; p = 2.50e-52). These findings suggest the training effectively enhanced provider understanding of systemic contributors to perinatal SUD and strengthened patient-centered, recovery-oriented approaches to care.

Following the training, participants reported statistically significant increases in comfort across all assessed clinical scenarios related to perinatal substance use (Table 4). The greatest improvements were seen in scenarios involving direct patient engagement and systems-level responses. For example, comfort with identifying local treatment options increased from a median of 2.0 [2.0, 3.0] to 3.0 [3.0, 4.0] (p = 4.15e-55), and arranging referrals to treatment programs improved from 2.0 [2.0, 3.0] to 3.0 [3.0, 4.0] (p = 1.67e-55). Participants also reported enhanced ability to initiate harm reduction conversations (pre: 2.0 [2.0, 3.0]; post: 3.0 [3.0, 4.0]; p = 1.56e-50) and intervene when witnessing stigmatizing language or behavior among colleagues (e.g., p = 5.95e-49 and 1.19e-46 for two peer-facing items). These findings suggest the training, beyond improving understanding or reducing stigma, directly contributed to improvements in provision of evidence-based care for patients.

**Table 4.**
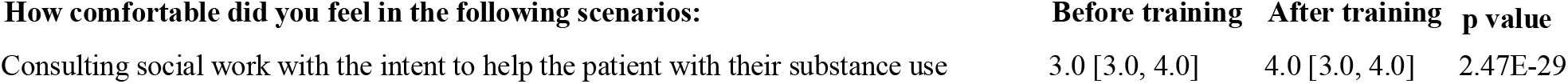
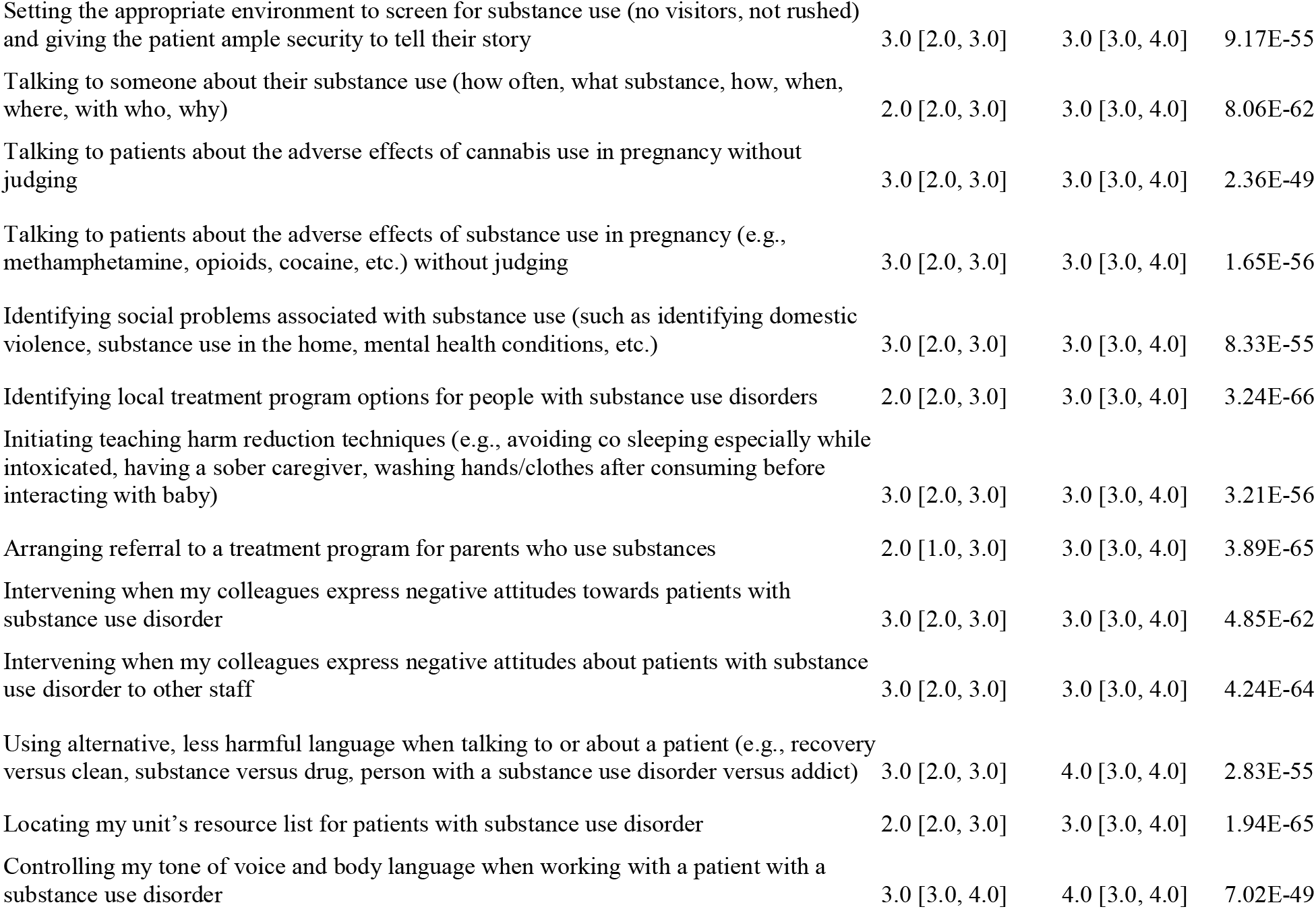
Comfortability with perinatal SUD.

Although some domains had relatively high baseline scores, such as consulting social work (3.0 [3.0, 4.0]) and controlling tone and body language (3.0 [3.0, 4.0]), all scenarios showed meaningful gains. Notably, participants felt more confident using non-stigmatizing language (pre: 3.0 [2.0, 3.0]; post: 4.0 [3.0, 4.0]; p = 1.78e-48) and setting appropriate conditions for substance use screening (pre: 3.0 [2.0, 3.0]; post: 3.0 [3.0, 4.0]; p = 2.48e-49). These results suggest that the training effectively improved provider comfort with both interpersonal and systemic components of perinatal SUD care.

Participants reported high levels of agreement with statements reflecting increased knowledge and confidence following the training (Table 5). The median response regarding increased understanding of treating patients with substance use disorders was 4.0 [IQR: 4.0, 4.0], indicating uniformly high perceived knowledge gain. Participants also reported increased confidence identifying appropriate resources and treatment options, with a median of 4.0 [3.0, 4.0]. Confidence in addressing stigma within the clinical environment was similarly strong, with participants reporting they felt more capable of intervening when colleagues made disparaging comments about patients with substance use disorders (4.0 [3.0, 4.0]). Participants showed a strong intention to share the information learned with colleagues and partners (4.0 [4.0, 4.0]), suggesting high potential for broader dissemination of the training’s impact.

**Table 5.** Overview of the training itself.

| AS A RESULT OF TODAY'S TRAINING, how much do you agree with the following statements? | Median [IQR] |
| --- | --- |
| My knowledge in treating patients with substance use disorders has increased | 4 [4-4] |
| I feel more confident identifying resources and treatment modalities for patients with substance use disorders | 4 [3-4] |
| I feel more confident intervening when I hear colleagues make disparaging comments about patients with substance use disorders | 4 [3-4] |
| I will share information learned with colleagues and partners for improved patient care | 4 [4-4] |

A subanalysis was conducted to compare the effectiveness of virtual versus in-person training formats. Across all 14 post-training clinical comfort items, participants reported similar levels of comfort regardless of training modality (Table 6).

**Table 6.**

| **How comfortable did you feel in the following scenarios:** | Virtual | In Person | p value |
| --- | --- | --- | --- |
| Consulting social work with the intent to help the patient with their substance use | 4 | 4 | 0.14 |
| Setting the appropriate environment to screen for substance use (no visitors, not rushed and giving the patient ample security to tell their story) | 3 | 3 | 0.34 |
| Talking to someone about their substance use (how often, what substance, how, when, where, with who, why) | 3 | 3 | 0.06 |
| Talking to patients about the adverse effects of cannabis use in pregnancy without judging | 3 | 3 | 0.67 |
| Talking to patients about the adverse effects of substance use in pregnancy (e.g., methamphetamine, opioids, cocaine, etc.) without judging | 3 | 3 | 0.74 |
| Identifying social problems associated with substance use (such as identifying domestic violence, substance use in the home, mental health conditions, etc.) | 3 | 3 | 0.72 |
| Identifying local treatment program options for people with substance use disorders | 3 | 3 | 0.01 |
| Initiating teaching harm reduction techniques (e.g., avoiding co sleeping especially while intoxicated, having a sober caregiver, washing hands/clothes after consuming before interacting with baby) | 4 | 3 | 0.20 |
| Arranging referral to a treatment program for parents who use substances | 3 | 3 | 0.01 |
| Intervening when my colleagues express negative attitudes towards patients with substance use disorder | 3 | 3 | 0.27 |
| Intervening when my colleagues express negative attitudes about patients with substance use disorder to other staff | 3 | 3 | 0.28 |
| Using alternative, less harmful language when talking to or about a patient (e.g., recovery versus clean, substance versus drug, person with a substance use disorder versus addict) | 4 | 4 | 0.31 |
| Locating my unit's resource list for patients with substance use disorder | 3 | 3 | 0.06 |
| Controlling my tone of voice and body language when working with a patient with a substance use disorder | 4 | 4 | 0.17 |

Median scores were largely consistent between groups, typically ranging from 3.0 to 4.0. No statistically significant differences were observed between virtual and in-person respondents on any individual item, as assessed by Wilcoxon rank-sum (Mann-Whitney U) tests (p > 0.05 for all comparisons). These findings suggest that the training was equally effective in improving clinical comfort with perinatal substance use scenarios whether delivered in person or virtually.

## Qualitative analysis

### What new insights were revealed as a direct result of the interactions with lived experience experts?

A total of 317 participants responded to the prompt, offering rich reflections on how hearing directly from lived experience experts shaped their understanding of perinatal substance use. The most frequently reported insight was a renewed sense of empathy and humanization, cited by 77 respondents. Many shared that the stories helped them see patients with SUD not just through the lens of diagnosis, but as individuals deserving of compassion and dignity. One participant reflected, “*More emphasis on empathy. Helps see a story behind a patient and understand their perspective*,” while another wrote, “*They are humans and deserve love and understanding just like anyone else*.” A second major theme, reported by 49 participants, involved the development of new communication skills. Respondents described learning how to open conversations in more supportive, nonjudgmental ways. One attendee noted, “*I was shocked to learn that patients might not answer truthfully if they sense judgment*,” while another emphasized, “*How to ask about substance use in a way that creates safety*.”

Another 48 participants gained awareness of the systemic barriers faced by patients, including stigma in healthcare, mistreatment, and under-recognized pain. “*Hearing how they were treated in hospitals made me reflect on how even small things can cause harm*,” one participant noted. Others reflected on the importance of ensuring timely, equitable care and empowering patients through available resources. A notable 26 responses described the experience as transformational, using words like “*eye opening*,” “*humbling*,” and “*practice changing*.” As one participant shared, “*It changed how I will approach patients in the future*.” Another stated, “*I come from this background—this session gave me strength*.” Lastly, 17 participants described new sensitivity to language and tone, highlighting the importance of avoiding stigmatizing terms like “clean” drug test results versus the less stigmatizing term of “negative” drug test results, and using recovery-centered, person-first language. One response summarized this shift well: “*Being more aware how your tone, body language, and words can help or hurt someone*.” Together, these reflections underscore that the inclusion of lived experience experts was not only impactful but essential to the training’s effectiveness. Participants left with greater empathy, sharper communication tools, and a deeper understanding of the barriers their patients face.

### How do you see peer coaching benefitting your future patients / medical community?

A total of 305 participants responded to the question, offering overwhelmingly positive reflections on the potential of peer coaching in clinical care,, which involves pairing a lived experience expert with a patient with perinatal SUD in order to provide patient support and advocacy. The most commonly cited benefit was the way peer coaching fosters trust and empathy between patient and the broader healthcare system, with 113 participants describing how patients may feel safer and more understood when supported by someone with lived experience. One participant shared, “*Peer coaching would be such an amazing tool for new moms with substance use disorder to feel safe and heard*,” while another emphasized, “*Patients may be more open when talking with someone who truly understands*.”

A second prominent theme, reflected in 94 responses, highlighted the power of lived experience insight. Respondents explained that peer coaches offer a form of relatability and credibility that providers alone may not be able to achieve. “*They’ve been there. That makes all the difference*,” one participant wrote. Others echoed this, noting, “*Patients listen to people who know what they’ve lived through*,” and “*Someone who’s walked through recovery brings a whole different perspective*.”

Participants also described how peer coaches could support care at a systems level, with 81 respondents envisioning peer integration across units and stages of care. Responses included statements like, “*Learning about different resources at* [hospital] *offers with peer coaching… made me realize how helpful they could be in postpartum*,” and “*They could be part of discharge planning, NICU follow-up, and prenatal care*.” Several participants noted the role of peer coaching in reducing stigma within clinical environments. One wrote simply, “*Better compassionate care*,” while another reflected, “*Peer coaches could help reduce stigma and change how staff talk about these patients*.” These perspectives suggest that peer presence may improve not only patient experience but also provider attitudes and unit culture. Finally, a smaller but meaningful theme emerged around communication bridging, with 7 participants emphasizing that peer coaches could help translate or soften clinical messages. One participant shared, “*I would love a translator to help patients understand that we are here to help*,” and another wrote, “*Sometimes patients are too scared to talk to us. A peer coach could bridge that gap*.” Together, these responses reveal that peer coaching is widely viewed as a promising strategy for improving trust, humanizing care, and creating more inclusive clinical systems for patients affected by substance use.

### Do you want more interaction with lived experience experts like the CU CON Recovery Coach Doulas or HardBeauty?

Responses to the question “Do you want more interaction with lived experience experts like the CU CON Recovery Coach Doulas or HardBeauty?” revealed strong enthusiasm for continued engagement, with the vast majority of participants responding affirmatively. Many described the interaction as one of the most impactful aspects of the training. Phrases like *“Yes! This was my favorite part,”, “YES!! More therapeutic environment!”* and *“Yes, I think recovery coach doulas would really help bridge a gap and benefit moms,”* reflected deep appreciation for the insight and relatability offered by individuals with lived experience of perinatal SUD. Several respondents emphasized that these interactions made the training more real and meaningful: *“This was an awesome training… the lived experience experts sharing their stories really helped me see their perspectives and helped me get a better understanding of how I can help*.*”* Another participant shared, *“They bring an insight that cannot just be read about in a medical journal*.*”* A notable number of responses also described how lived experience could be applied directly in clinical practice, suggesting roles such as bedside advocates, intrapartum educators, or resources for patients postpartum. One participant suggested, *“It would be so sweet to have recovery coach doulas in deliveries to advocate and also to educate nurses and providers,”* while another proposed using *“iPads to connect patients to peer supports*.*”* Others called for even more time with these speakers, with one person noting, *“I wish I could have heard more speakers,”* and another stating, *“More time to ask them questions would be great*.*”*

A smaller subset of participants offered neutral or conditional responses, with comments such as *“Maybe,” “Sure, but it would be helpful to know how to help moms in the NICU,”* or *“Yes - if they are able to give us action items we can implement*.*”* A few respondents were not interested or unsure about additional interaction, with responses like *“No thank you,” “Not at this time,”* or *“I have worked with SUD patients and feel I already have a deep understanding*.*”* However, even among these comments, many framed their response in terms of role clarity or existing familiarity, rather than opposition to peer involvement. Overall, participants overwhelmingly affirmed that interactions with recovery coach doulas and lived experience experts enriched their understanding of substance use disorders, helped humanize their clinical care, and would be a welcome and valuable component of ongoing training and patient support.

### Is there anything else you would like us to know?

In response to the open-ended prompt “Is there anything else you would like us to know?”, participants provided candid, heartfelt feedback that spanned appreciation, requests for clarification, and critical reflections. Many respondents praised the training as “*powerful*,” “*impactful*,” and “*eye opening*,” with numerous comments highlighting the value of integrating real stories from individuals in recovery from SUD. Several described this as the most meaningful component, expressing a desire for more time for Q&A with lived experience experts and future trainings that continue to prioritize personal narratives. One respondent said “*going into this training I was frustrated that I was required to go to another training, by the end I was thankful*.”

## Discussion

This study demonstrates the effectiveness of a brief, structured training program in reducing stigma and improving clinical confidence among healthcare providers who care for pregnant and postpartum individuals with substance use disorders (SUD). The integration of lived experience experts, paired with interactive discussion and trauma-informed education, meaningfully enhanced the training’s impact. Participants showed statistically significant improvements across all measured domains, including knowledge, empathy, and clinical comfort, and open-ended feedback underscored the emotional resonance and practical utility of hearing directly from individuals in recovery. The program not only addressed persistent misconceptions (e.g., that recovery depends solely on willpower), but also provided participants with concrete, patient-centered strategies for nonjudgmental communication, stigma intervention, and linkage to treatment.

The need for such interventions is clear. Pre-training assessments revealed enduring provider stigma, discomfort when engaging with perinatal patients who use substances, and limited awareness of local treatment and harm reduction resources. These challenges were reported across roles and units but were especially pronounced in settings such as the NICU and among night-shift staff, groups that often feel excluded from formal care planning and may lack access to team-based support. The training responded to these gaps by equipping staff with clear communication tools, emphasizing interdisciplinary collaboration, and reinforcing a harm-reduction framework that meets patients where they are.

Importantly, this training model differs from traditional educational formats. Prior research has shown that asynchronous, didactic modules often lead to limited or short-lived improvements in provider attitudes toward patients with SUD.^7-14^ In contrast, this program’s emphasis on lived experience, storytelling, and active reflection produced deeper emotional engagement and more durable learning. Qualitative findings suggest that participants internalized these lessons in tangible ways, reporting changes not only in knowledge, but also in body language, tone of voice, and word choice. These behavioral shifts, while self-reported, point to meaningful changes in clinical practice that may not emerge from more passive forms of training.

Another key strength of this program is its scalability. The intervention was brief (90 minutes), offered in both virtual and in-person formats, and reached providers across diverse roles, units, and geographic settings. Subanalysis showed no statistically significant differences in outcomes between virtual and in-person participants, suggesting the training can be flexibly delivered without compromising efficacy. This adaptability is critical for broader implementation, particularly in under-resourced or rural settings where in-person sessions may be less feasible.

Despite these strengths, several limitations warrant consideration. Participation in the survey was voluntary and limited to those who completed at least 80% of the evaluation, raising the possibility of self-selection bias. Because demographic data were only collected from survey completers, we could not assess whether non-responders differed meaningfully from responders. The training was a single-session intervention; although short-term gains were evident, long-term impact remains unknown. However, a six-month follow-up survey is currently underway to assess retention of knowledge and sustained changes in clinical attitudes. All findings were self-reported, and future research should incorporate objective outcome measures such as chart reviews, patient feedback, or clinical audit data. Additionally, some qualitative feedback may reflect social desirability bias, particularly in the context of highly stigmatized topics such as SUD and provider bias.

Nonetheless, this study contributes to a growing body of evidence supporting the integration of lived experience into provider education. When designed with authenticity, interactivity, and community partnership, short trainings can shift not only individual provider attitudes but also broader unit culture. Hospitals and clinical teams are encouraged to adopt or adapt similar models as part of ongoing staff development. Embedding recovery voices, small-group discussion, and trauma-informed frameworks into routine training can strengthen provider confidence, reduce stigma, and advance more equitable, compassionate, and effective care for families affected by perinatal substance use.

## Data Availability

Data will be made available upon reasonable request to the authors.

